# Assessment of Learning Organization Practice and Disciplines in Case of Sire District Public Health Office, Arsi Zone, Oromia Region, Ethiopia, 2018:Qualitative Case Study

**DOI:** 10.1101/2020.10.02.20205807

**Authors:** Biniam Worku Hailu

## Abstract

Learning Organizations are organizations where people continually expand their capacity to create the results they truly desire, where new and expansive patterns of thinking are nurtured, where collective aspirations are set free and where people are continually learning how to learn together. The aim of this study was to assess Learning organization practice and disciplines in case of Sire District Public health office. Learning organization if implemented in the health institutions it enables ignition of newly created more another’s learning organization units moreover, learning organization concept if implemented in the organization it can enables organization to achieve their goal systematically besides this, it also enhance employees their job commitment whereas, the organization not implemented learning organization concept might be opposite of this or vice versa. The study employed qualitative case-study design in sire District public health office. Document review, in depth interview and Focus group discussion with flexible probing techniques were employed to collect the data from 3 key informants and 2 FGD from January 1, 2018 to January 11, 2018. A semi structured interview guide was prepared to explore key informants’ idea about current situation and efforts made to enable organization to be a learning organization and also thematic analyses were used to analyze the findings of the study. The study result revealed that the organization assessed in both parameter of learning organization (five disciplines Peter Senge and Pedler et al criteria of practice of learning organization) the organization was practiced preponderance of learning organization concept was practiced within five disciplines of learning organizations there were implemented in their office. The finding of the study revealed that sire district public health office was practiced learning organization and disciplines in there organization that was exercised as good and the organization also participate entire employees and stakeholders to carry out organizational plan that were align with national strategies of the country moreover, the organization had created enabling learning opportunities to enhance a learning organization among themselves and others.

## 1. Introduction

A health system consists of all organizations, people and actions whose primary intent is to promote, restore or maintain health according to the World Health Organization WHO [1]. Healthcare organizations are composed of health care professionals from multiple disciplines forming several interconnected care teams that strive to provide safe and consistent care [2].The care teams have to coordinate and communicate amongst their team members and with other teams to function in a cohesive manner to execute their task the highly coordinated to satisfy patient care and then to achieve organizational goals [3].

Learning Organizations are organizations where people continually expand their capacity to create the results they truly desire, where new and expansive patterns of thinking are nurtured, where collective aspirations are set free and where people are continually learning how to learn together [4].

The important component of the definitions above is the requirement that change occur in the way work gets done. Learning in an organization means the continuous testing of experience, and the transformation of that experience into knowledge-accessible to the whole organization, and relevant to its core purpose [5].

According to Federal Democratic Republic of Ethiopian government, Minister of health in their Transformation plan, stated that best practice and experience should be disseminated amongst health facilities. In this context the health sector transformation plan incorporate four basic pillars; amongst those pillars District transformation is one of its pillar. The District transformation in the context of health sector indicates the concept relate with creation of learning organization and this process pass through selecting District those who have best experience and integrated activities (Learning organization) and disseminating best experience to other health facilities (Organizational learning). This leads to transformation of organizational learning to learning organization [6, 7].

The study aimed to assess learning organization practice and disciplines in case of sire District Public Health office. Furthermore, this study will also generate knowledge that can contribute to ongoing Research efforts and also will be used as additional input for further study in the same area of inquiry.

### Operational Definition of Learning Organization

Learning Organizations: are organizations where people continually expand their capacity to create the results they truly desire, where new and expansive patterns of thinking are nurtured, where collective aspirations are set free and where people are continually learning how to learn together [8].

Personal mastery-Learning to expand personal capacity to create results most desire and creating an organizational environment which encourages all its members to develop themselves toward goals and purposes they choose [8].

Mental Models-reflecting upon, continually clarifying, and improving internal pictures of the world, and seeing how they shape our actions and decisions [8].

Shared Vision-Building a sense of commitment in a group [8].

Team Learning-Transforming conversational and collective thinking skill [8].

Systems Thinking-a way of thinking about, and a language for describing and understanding forces and interrelationships that shapes the behavior of systems [8].

## 2. Methods and Materials

### 2.1. Study Area and Period

The study was conducted in Sire District from January, 1-11, 2018. The district is one of the 26 districts found in Arsi zone. The district has 18 kebeles, 1 urban kebeles and 17 rural kebeles. The district is located 145KM and 75 KM from A.A and Asela respectively. The population of the district was 73,970 in 2007 according to National census reported the rural population accounts for about 67,757 (91.6%) of the population [9]. The district has four health centers and 17 health posts.

### 2.2. Study Design

Case study (Qualitative) design was employed

### 2.3. Population

#### 2.3.1. Source Population

All Health Workers Working in Sire District Public Health office were source of population.

#### 2.3.2. Study Population

District Health office Head, vice head, District Health Office process owners, Primary health care directors and kebele’s health extension workers.

### 2.4. Sample Size

A total of two Focus group discussion (with minimum and maximum duration of 57 and 82 minutes respectively, and with the number of participants ranged from ten to twelve), three in-depth interview and Document reviews (1) operational and program plans;, changes to existing programs, or to end programs; (2) Performance appraisal result (3) organization-level documents (e.g., organizational chart, strategic plan). In total, 8 documents were submitted all available documents were reviewed).

### 2.5. Sampling Technique and Procedure

Purposive sampling technique was employed in selection of participants for both in-depth interview and focus group discussion.

The numbers of in-depth interview were determined by saturation of information. However, a total of three In-depth interviews were carried out in sire District health office. In-depth interview was conducted for 3 key informants consisting of; one health extension worker, 1 Primary health care unit director, 1 district health office Head who was selected purposively based on their expertise and experience within the district health office.

Focus group discussion was conducted in two groups that were purposively selected from Health Extension Workers, District health office Core process owners, And each group consisted of 12 and 10, experienced moderator facilitated the discussion using semi-structured interview questionnaire focusing on Peter Senge Five Discipline of learning organization and 8 Criteria of learning Pedler et al challenge to be learning organization and their contribution in solving the problems. The FGD took an average of 69 minute to discuss the issue under study.

During this study District Health office plan and achievement on Wash, health extension data concerning maternal and child health, nutrition and others program areas were reviewed.

### 2.6. Data Collection Techniques

In-depth interview, Focus group discussion, and Review of documents were used. A total of two FGDs were conducted with Health Extension Workers, district health office Core process owners and PHCU directors.

As well as FGD; in a way that appropriate time and comfortable place of meeting was selected and organized. Before conducting the discussion, explanation and elaboration of the need to do the FGD was made and the participants were asked for their willingness to participate in the FGD. Each FGD group was conducted by two team members composed of recorder and facilitators who were fluent speaker of local language.

In-depth interviews; were made with three key informants Sire district public health office it was made by who were fluent speaker of local language. Audio record and notes were used for taking qualitative data in FGD and in depth interview.

### 2.7. Data Collection Tools

Data were collected using FGD guiding questions, Document review checklist (data extraction tool) and In-depth Interview guides.

To assess sire district public health office whether the organization is learning organization or not the investigator had adopted two study tools for interview and FGD to make assessment coherent and enhance accuracy before the investigators had adapted a semi structured questionnaire from Pedler et al and five disciplines Peter Senge [10,11]. The determination whether the organization learning organization or not the assessment tools trace the following main points:

1. Assess the organization in terms of criteria (dimension) of learning organization
2. Assess the organization in terms of Discipline of learning organization?
3. Learning organization cycle
4. Challenge the organization faced to be learning organization

The jotted down above points address the key the concept of learning organization. the result of assessments tools are analyzed and presented similar idea amalgamated together have been described to make description clearly also comparison also made between assessment tools results vs. scholars view to make the study rational.

### 2.8. Data Analysis

Data were transcribed in to text by principal investigator after listening audio recoded and reading the written interview notes. Different ideas in the text are color coded and merged in their thematic areas and thematic analyses were employed manually. And then the analyzed data were presented in text, table and diagram.

### 2.9. Ethical Considerations

Formal letter of permission was obtained Sire Woreda public health office to communicate with Health Extension workers, Primary health care directors and district health office core process owners.

## 3. Result

In FGD open ended questions attached on the Annex guided the data to capture information towards 8 Criteria of learning organization Pedler et al and In depth semi-structure interview question guided Peter Senge Five Discipline of learning organization and challenge to be learning organization and their contribution in solving the problems [10, 11]. The data were organized and managed manually. Open coding assists us to identify specific categories of learning organization mentioned by discussants while axial coding used to connect Criteria learning organization assessment sited prior identified by discussants.

### 3.1. Focus Group Discussion (FGD)

Two FGDs (One FGD with Hews and one FGD with job process coordinators and expert were conducted. Each FGD comprised of ten to twelve participants. Participants of FGDs were purposely selected. A total of 21 participants (10 male and 12 females) were included. The age of the participants ranged from 20 to 54 years. All participants engaged well with the topic and responded enthusiastically to the questions.

During the qualitative data collection of FGD were learning organization assessments. Finally, the findings were generalized in to eight themes.

1. Adopt a learning approach to strategy
2. Participative policy-making
3. Access and transparency of information,
4. Formative accounting
5. Internal exchange and dialogue
6. Reward flexibility,
7. Inter-company learning,
8. Self-development opportunities for all

A learning organization is continually getting ‘smarter’ because learning is planned, systematic and in alignment with the organization’s strategic goals. In order to get smarter, the organization needs to capture its organizational knowledge. Pedler et al have described the process of how organizations learn, and identify the outcomes of the process as the development of core competencies, which are ‘… the collective learning in the organization’. This occurs at all levels and functions within the organization [10]. On the surface, developing core competencies has an intuitive appeal, examined in this literature when the investigators assessed sire District health office by criteria of learning organization to determine the organization is whether learning organization or not the investigators examined the sire District health office according to. Pedler et al scholar learning organization assessed. Data were transcribed in to text by principal investigator after listening audio recoded and reading the written FGD notes. Different ideas in the text are color coded and merged in their thematic areas and thematic analyses were employed manually. And then the analyzed data were presented in text. Thematic areas are as follows;

#### Theme 1. Organizational adopts a learning approach to strategy

In this study most of participants said that organization Adopt a learning approach to strategy and the organization also participate entire employees and stakeholders to carry out organizational plan that was also align with national strategy of the country. One of the discussant from FGD 2 stated that “……*when* ***S****ire District health office carried out its plan it involves the entire employees and important stakeholders. the District health office plan started from situational analysis after the plan is performed its achievement is monitored consistently through Performance monitoring team carry out its tasks according to plan they routinely identify root causes problems and develop action using problem investigation and action plan form this make the office to follow up and implementation proposed intervention were to identify early and make instance decisions for the gap manifested*”

#### Theme 2. Participation of employees and stakeholders

In this study most of participants said that the organization observe a problem from different aspects and encourage employees to engage across the organization and with outside environment to bring solutions together.

One of the discussant from FGD 1 stated that “……*The organization participate employees and stake holder starting from organizational plan up to achievement of the plan. The district health office oriented the employees regarding their job duties and also empowered to perform their job duties under full of accountability’’*

#### Theme 3. Access, transparency information

One of the discussant from FGD 1 stated that “……*The access of information in the organization is possible to be seen in two perspectives. The health office initially accessibility of information toward training concerning capacity building of the organization in this case the organization enable Entire employees has to took integrated and program training so as to make enhance accessibility training the organization to adopt various strategies, the first is implementation of on job training in the organization example with trainee staff after taking training either at zonal or regional level and when he/she return back to work place he/she expect to rendered on job training to concerned staff members and also human resource managers of organization audited training. Secondly, the organization staff including supportive staff and health workers access information regarding their job in the organization via one to five network on daily bases*’’.

Also another discussant from FGD 2 stated that “……to make information within the organization transparent; every department unit posted their performance in the office in the visible place. This enables possible everyone to saw their performance without support of the departmental personnel with shorter span of time and on the other hand transparency of information in the organization adhered by transparent financial system control such as the ledger balanced book put on visible place and also the liquidation and expenditure of the budget was posted monthly and quarterly in the office”.

#### Theme 4. Formative accounting system of organization

One of the discussant from FGD 1 stated that “…… The formative accounting system of the our organization Emphasis on the formative processes through which control procedures occurred and the tasks performed are then argued between the monitoring actor and the controlled one, with the purpose is to generate progress and learning. Constructive approaches should be shared on an organizational level and become part of the overall procedural system. The accounting, budgeting and reporting systems have to be set up so they promote learning.’’

#### Theme 5. Internal exchange and dialogue

In this study most of participants said internal exchange and dialogue system of the organization was decentralized system that mangers or team process owners didn’t make decision solely rather managers consider Dialogues open space for critical thinking and bring logical, appropriate and new ideas to contribute to the culture of learning for their organization.

One of the discussant from discussant from FGD 1 stated that *“……Internal exchange and dialogue system the organization is decentralized the manager does not solely make decision rather participate employees in the decision making. Employees also share information themselves daily through one to five network they also solve the problems and make decision in their network regarding to the function and responsibility which are articulated within the organization this communication and dialogue schemes make the communication flow of the organization relevant and crucial for the organization”*.

#### Theme 6. Reward flexibility

In this study most of participant said that the organization had facilitate reward and career benefit scheme for their employees depend on performance status employees. Possible explanation for this could be promoting a culture of engagement.

As an example one discussant from FGD 1 stated that *“……The organization rewarding systems depend on performance status employees. The District has organized team those who perform reward and career prospect the so called reward and career team. The team does their own work plan and also work aligned with performance appraisal committee*.”

One of the discussant from FGD 1 stated that *“……Sire District health office since 2014 its performance evaluation at Zonal and District level its Rank is first owing to this, the District Role models employees has got seven Master and Three Degree the employees got Tertiary education even though this education solely given to a few role module employees, it provide opportunities for other individual employees to have bright Career prospects. And also for the rest of workers the organization facilitate might be either extrinsic reward (promotion, increase in pay according to public service laws) or intrinsic reward (greater fulfillment through a more demanding or higher-status job)”*.

#### Theme 7. Inter-health facility learning

In this study most of participant said that the organization collected the best experience and share those best experiences for different health institution. In healthcare settings, the learning acquired from the new knowledge should be deep rooted and become part of daily operations of health institution this enables.to be Acquired knowledge and incorporated from outsiders embedded in the working systems, practices, and structures can be used and shared to improve performance

One of the discussant from FGD 1 stated that *“……the organization initially start collecting best experience via photograph, video and report. The organization organize the collected data of best experience of primary health care unit share those best experience for the rest primary health care unit during review meeting conducted afterword the District facilitate the primary health care unit to make action plan to implement and improve those best practice through the process of above indicated jotted down stage the District reached learning organization level owing to this the regional as well as the zonal health office selected sire District health as pilot learning organization center and the District also share experience for different District health office, health facility and zonal health department”*

#### Theme 8. Self-development opportunities for all

One of the discussant from FGD 1 stated that *“……There were a lot of opportunities, materials, and resources available for learning. Thus resource has been allocated by the government and Non-Governmental organization such as transform health care project which facilitate break through grant and work on transformation agenda for enhancement of capacity of employees focuses on the possibility to access learning opportunities and to start personalized development processes but, the allocated resource for us not adequate but, we use it as intensively*” This criterion could be related to Senge’s discipline of personal mastery and with Stewart’s requirement of individuals committed to self-development. The problem here consists in who owns the learning, the employees or the employers, and the uses to which the new learning will be put.

### 3.2. In Depth Interview

A total of three key informants were planned to in depth interview from three planned key informants three of them were accessed with minimum and maximum duration of 50 and 95 minutes of interview respectively. Out of the three interviewed key-informants, one was second and two were first degree holders. Two of them were males and one was female. By the position they held, one were deputy or vice heads, one PHCU Director and one job process coordinator. The age of the participants ranged From 23 to 48 years. In-depth interview results were merged and presented together by quoting the words of the interviewees.

During in depth interview data collection were learning organization assessment finally, the findings were generalized in to five themes.

Theme 1 …Personal mastery

Theme 2. Mental Models

Theme 3 Team Learning

Theme 4. Shared Vision Theme

Theme 5. Systems thinking

**Table 1.**
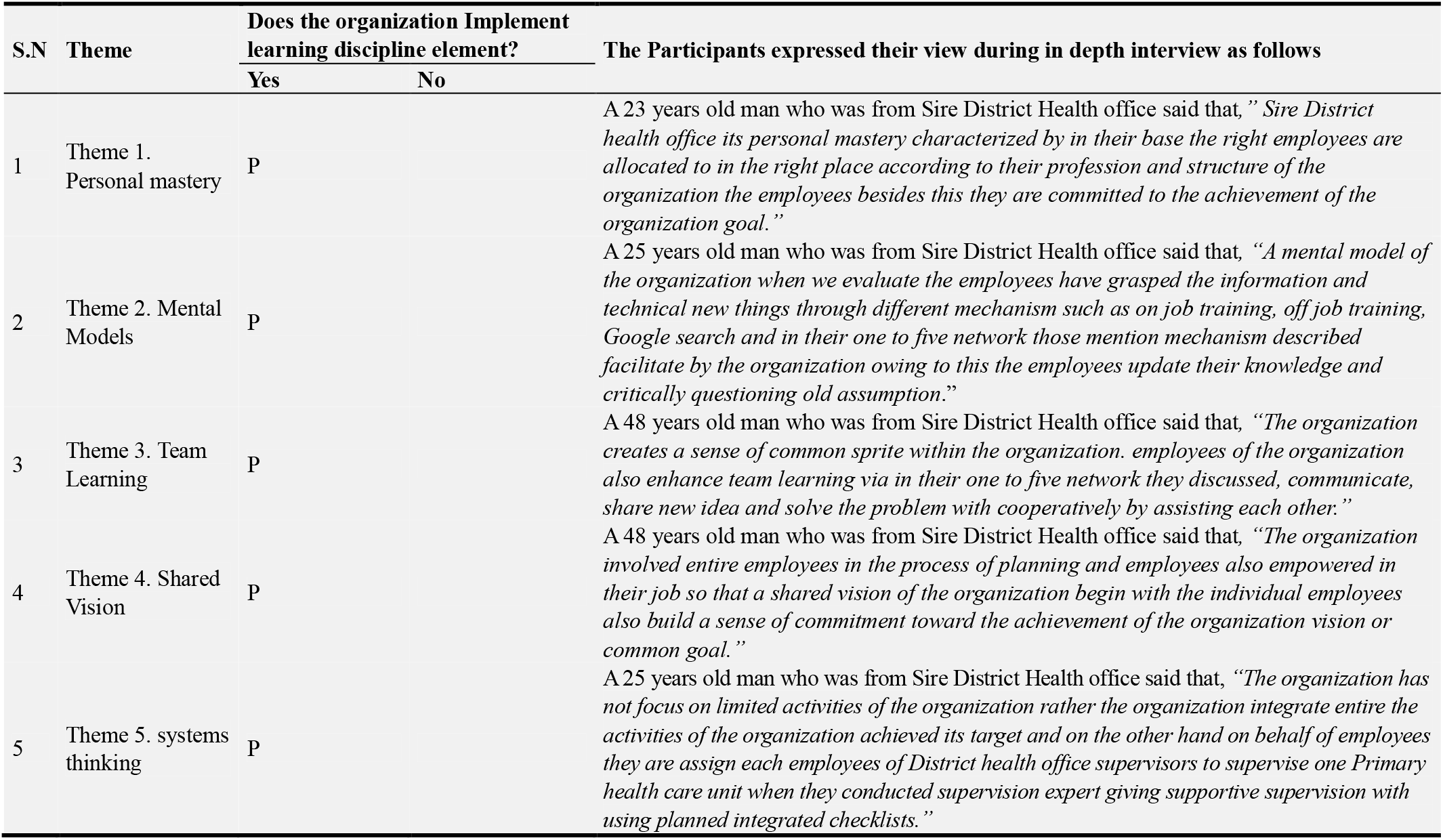
The five Disciplines of learning organization assessment of sire District health office, 2018.

### 3.3. Document Review

The purpose of the document review was to examine if/how the concept and approaches of learning organization were incorporated in the organization’s written items and to discern changes in the presence of evidence and over time. representatives were asked to provide a sample of recent documents, including: (1) operational and program plans, changes to existing programs, or to end programs; (2) Performance appraisal criteria result; (3) organization-level documents (e.g., organizational chart, strategic plan). In total, 8 documents were submitted all available documents were reviewed the documents result regarding to learning organization the organization phase of being learning organization as shown below summarized figure 1.

**Figure 1.**
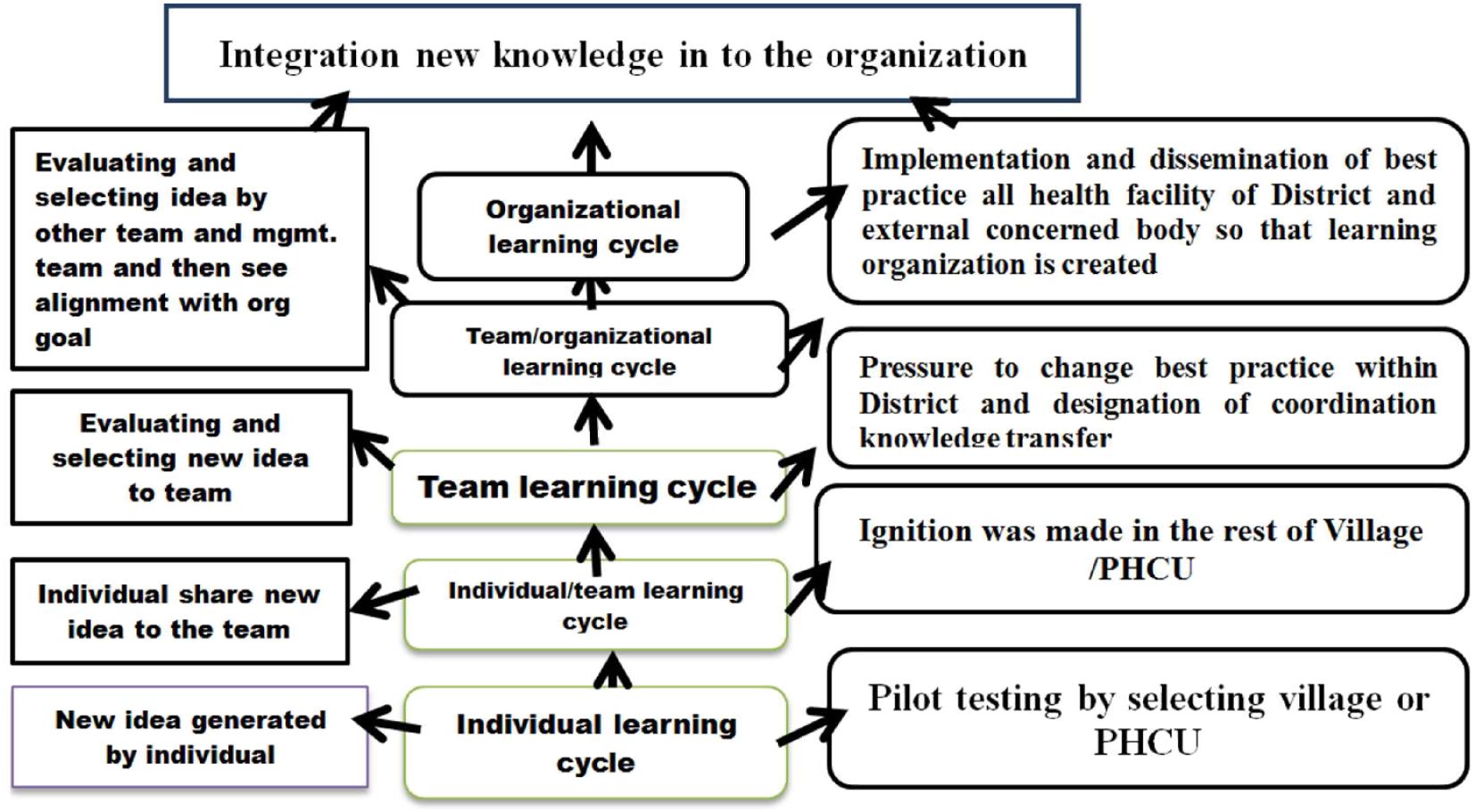
The sire District learning organizations cycle its implementation status, 2018.

This figure shows clearly learning organization cycle of sire District public health office Learning Organization cycle of the district health office which indicate is total system or integration of knowledge into the organization. The Organization learning cycles are not merely collections of individuals, yet there are no organizations without such collections. Yet the organizations learn only through the experience and learning of individuals. It is clear that a major challenge is that of transforming or transferring individual learning into organizational learning. Learning takes account of the creation and diffusion of knowledge at various levels: the team, departments, and organization.

The document review analyzed from sire District Public health office specially the District plan vs. performance, number of model employees having high Performance appraisals grade had got recognition letters from different higher officials and also on behalf of the organization got different reward from different organization based on good performance in District as well as at zonal and regional level.

## 4. Discussion

The output of frame learning organization this study was analyzed and conceptually connected The results revealed that majority of discussants raised sire District public health office as found as learning organization in both parameters five “learning disciplines and eight Criteria of learning organization Pedler et al [10, 11].

The sire District public health office status in both the FGD and in-depth interview the study result revealed that it was a benchmark District for horizontally or vertically inter related organizations or institution. According to FGD result with the eight learning organization criteria/parameter like adopt a learning approach to strategy, In Generative Learning Organizations, Participative policy-making access and transparency of information, formative accounting, internal exchange and dialogue reward flexibility, Inter-company learning, Self-development [10]. were accordingly implemented in the District health office level up to the health facilities linked in primary health care unit. when this result compared with study conducted in Suffolk University, Boston, Massachusetts, USA on assessing change: can organizational learning “work” for schools which revealed that if schools were not be a learning organizations it’s difficult to improved organizational performance as well as minimizing turnover [12].On the other hand the study conducted in Kenya revealed that specific knowledge scheme exerts competitive advantage for the organization [13]. This finding was in lined when compared with learning organization criteria implementation sire District as it was an exemplary public organization that have nice performer as knowledge and experience sharing organization. When Senge’s five principles of learning organization assessed qualitatively the five principles in sire District were well developed when compared to study done in Technological Educational Institution of Western Greece that are developed well shared vision, team learning and system thinking only out of the five learning organization principles[12]. The result analyzed with clear thematazed from the FGD and in-depth interview discussants told as to became as learning organization as an organization many challenges was passed through. The five senge’s learning principles was fitted as an organization and the document review analyzed from sire District specially the District plan performance, number of model workers with individually having high grade work evaluation and recognition letters took from different higher officials suggested that those principals are well and integrally implemented.

Finally, findings from the document review, in depth interview and FGD were well analyzed and give sense that the studied organization was learning and other might be used it as a bench mark.

This study has limitations of raising the same idea and influenced by others idea during FGD discussion as well as there might be self-reporting bias during in-depth interview. Another limitation of this study was limited literatures to discuss in Africa and Ethiopia context.

## 5. Conclusion

The finding of the study revealed that sire district public health office was practiced a learning organization and disciplines concept in there organization that was embraced as good and the organization also participate entire employees and stakeholders to carry out organizational plan that was also align with national strategies of the country moreover, the organization had created enabling learning opportunities to enhance a learning organization among themselves and others.

## Data Availability

all data are available

## Consent for Publication

I agree with the publication of this manuscript

## Acknowledgements

My sincere gratitude should goes to Sire woreda health office for their holistic cooperation for this study, In addition I would like to thank the study participants for their voluntary participation and finally I would like to thank data collectors for their cooperation throughout data collection process.

